# Super spreader cohorts and covid-19

**DOI:** 10.1101/2020.05.15.20103184

**Authors:** Patrick Van Esch

## Abstract

A simple two-cohort SIR like model can explain the qualitative behaviour of the logarithmic derivative estimations of the covid-19 epidemic evolution as observed in several countries. The model consists of a general population in which the *R*_0_ value is slightly below 1, but in which a super-spreading small subgroup with high *R*_0_, coupled to the general population, is contaminating a significant fraction of the population. The epidemic starts to slow down when herd immunity is reached in this subgroup. The dynamics of this system is quite robust against non-pharmaceutical measures.

## 1 Introduction

The covid-19 epidemic dynamics has to be understood in order to have an intelligent policy that optimizes society’s well-being. Without understanding that dynamics, one might take measures that are highly ineffective and/or place a huge burden on society, with a cost-benefit ratio that is far from optimal.

Unfortunately, not much high-quality data is available that allows us to test against sophisticated epidemiological models. The most important data, which describes the propagation of infections, is simply not publicly available for most populations. Confirmed cases depend strongly on ever changing policies of testing and are potentially severely biased. They are very bad proxies to estimate infections. Hospitalisation entries are also depending on decisions which are different from country to country, and may be changing over time.

The only more or less reliable data are the numbers of dead, which are most probably within a factor 2 or 3 from the actual numbers of victims of covid-19. The problem with this proxy is that there’s a long delay and a large smoothing in time, due to the probability distribution that links the event of being contaminated with dying of the disease. The simple SIR model suggests that one can obtain an estimate of the *β* − *γ* factor by calculating the deconvolved logarithmic derivative of a proxy curve for contamination.

In a recent paper [3], the logarithmic derivative of the curves of the number of deceased people of different countries has been calculated, for countries that have applied different sets of non-pharmaceutical policies, and have different climates. The behaviour of this logarithmic derivative seems to be quite universal, independent of the measures taken, and of other specific factors. The universal curve is a more or less linear decrease of the logarithmic derivative, starting around 0.3 per day, and steadily decreasing to a value of about –0.05, and then remains more or less constant. This might be an effect of the policies put in place, but it might also be the natural dynamics of the epidemic, independent of any policy.

We re-calculated the same kind of curve here, for a few selected European countries. We took the data from [1], and we applied a moving 7-day average to have less noisy data, and remove weekly cyclic phenomena. Calculating the logarithmic derivative of the reported deceased cases for some European countries, we confirm the behavioural findings of the cited paper:

**Figure 1:**
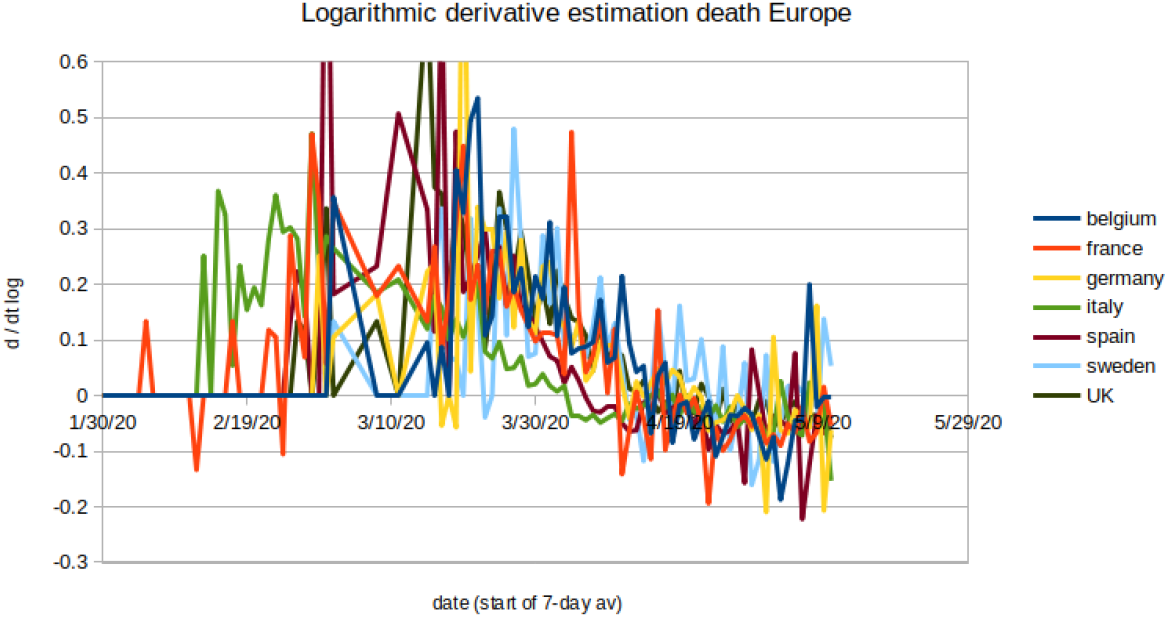
Logarithmic derivative of the weekly smoothed covid-19 deaths for some European countries.

We note the extreme similarity between all these curves. Only Italy has a somewhat slower decay, but it starts out also around 0.3. All these curves level off towards the same slightly negative value of about –0.05. It is also intriguing that the tendency sets in quite a few weeks before one expects to see the effects on the deaths of the lock-down policies put in place in some countries. It was the essential point raised in [3]. This doesn’t necessarily mean that those policies have no effect; it means that the linear decrease is not much affected by those policies. We will come to that.

A more interesting curve to look to is that of Brazil. Brazil is interesting for several reasons. The first one is that the country is in the Southern hemisphere. All seasonal effects should be opposite there. The second one is that the policies put in place are quite different from those in Europe (only very partial lock-downs).

We notice again the same behaviour. However, it seems that the “constant part” is rather slightly positive rather than negative even though the data is noisy and it is not clear if there’s still not a slope downward.

**Figure 2:**
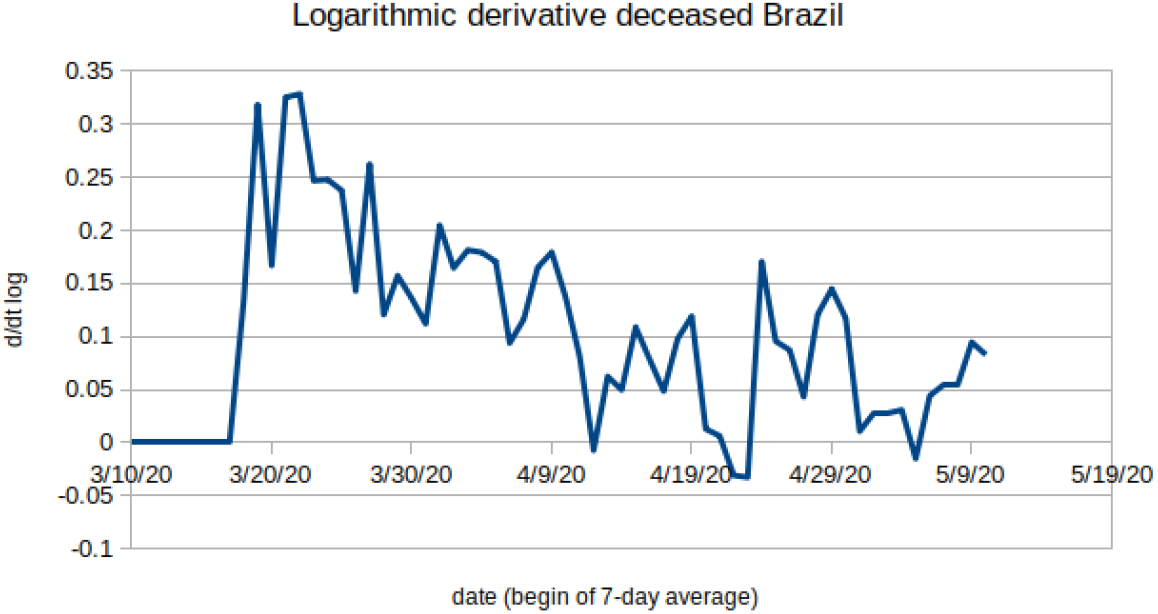
Logarithmic derivative of the weekly smoothed covid-19 deaths for Brazil.

We find again a similar behaviour by analysing for instance, the entries in Belgian hospitals since March 15. These data were taken from [2]. Unfortunately, there is no data available before this date, so the plot is truncated on the left. We had to average the entries over a week because there were weekly periodic effects and the daily data was too noisy, but if one averages over 7 days, and one calculates the logarithmic derivative of the hospital entries, one finds again the same universal curve:

**Figure 3:**
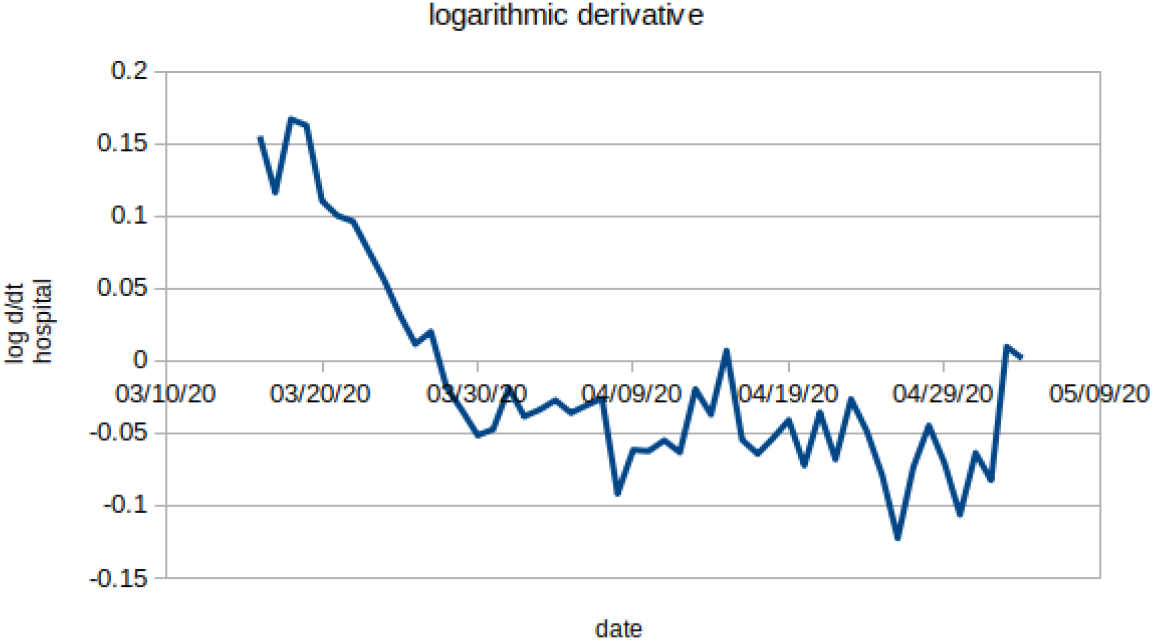
Logarithmic derivative of the weekly smoothed hospital entries for covid-19 cases in Belgium.

In this paper, we try to build a model that displays a similar qualitative behaviour. We do not fit that model to any real-world data, and we do not pretend at all to have a working and predictive model for the covid-19 epidemic. We are only exploring a potential mechanism that might explain the universal qualitative behaviour of the logarithmic derivative of observed data by building a very simple model exhibiting a similar qualitative behaviour.

## 2 Models

### 2.1 The classical SIR model

In the classical SIR model, there are 3 compartments: the susceptible population, the infected and contagious population, and the recovering and non-susceptible/noncontagious population. In reality we only care about the first two compartments, because we want to study the dynamics of contagion, not of recovery.

The SIR model models the contagion dynamics with two hypotheses. The first is that any member of the I compartment will potentially contaminate *β* people per day, but of these *β* potential people, only a fraction *S/N*_0_ is actually susceptible to be infected. The second is that each member of I has a probability of *γ* per day to become a member of R (that is to say, to lose its contagious statute, and to be removed from the people concerned in the contagion process). Essentially, it comes down to saying that on average, a contagious person is contagious for about 1/*γ* days.

The equations implementing the above hypotheses are as follows:

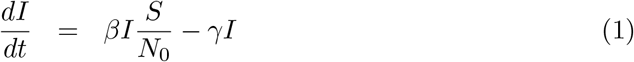

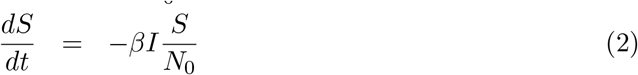

The logarithmic derivative of the infected cases, or of any proxy that is proportional to the infected cases (such as the number of dead) equals:

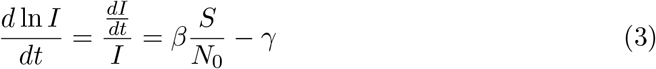

We recall that *R*_0_ is defined to be:

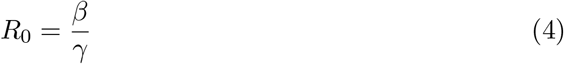

and is interpreted as being the average number of people that a contagious person will contaminate over time, if the population is naive (that is, *S* = *N*_0_). If *R*_0_ > 1, the illness can start propagating in a naive population.

In the same vain, *R*(*t*) is defined as:

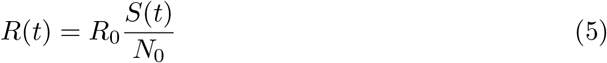

and is interpreted as being the instantaneous number of people a contagious person will contaminate over time in a population that has already some immunity. If *R >* 1, the illness continues to propagate, if *R* < 1, the epidemic starts dying out. The condition *R* = 1 equals the condition 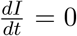 and is the point where the number of contaminated people starts diminishing (when the epidemic starts dying out). It is not the point where no more new infections are happening, but it is the point where herd immunity has been reached. If at that point, there are still a lot of contagious people, they will continue for a while to contaminate others, but their numbers will decrease.

Herd immunity is reached when:

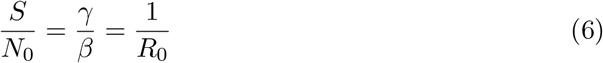

in other words, when a fraction 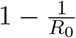 of the population got ill.

In an SIR model, initially the logarithmic derivative equals *β* − *γ*, and, as the fraction of the susceptible population *S/N*_0_ decreases (as “immunity” increases), the value evolves towards an asymptotic value between 0 and −*γ*. When the epidemic ends, the logarithmic derivative tends towards a constant.

At first sight, this corresponds exactly to what has been observed for the covid-19 epidemic proxies: a steady almost linear decrease from a value around 0.3 towards a value close to –0.05 or so, followed by a constant value. However, that would mean that “herd immunity” has been reached in those populations. Most preliminary studies seem to indicate that in most countries, herd immunity is still far away, and only about 10% or so of the population has been removed from the “susceptible” status.

### 2.2 Super spreader sub population model

In order to try to reconcile the behaviour of the logarithmic derivative which points to “herd immunity reached” and the low attack rate in the overall population, we propose to consider a small super-spreader subgroup in the population.

We consider a small group which is initially also in a susceptible state, and which will also evolve into its own infectious state, and end up in a recovering state. However, we suppose that this small group can infect also a significant amount of people in the general population. The index 1 is used for the general population (of size *N*_0_) and the index 2 is used for our super spreader group (of size *N_S_*).

If we were to have two separate groups which do not interact, but of which the second group has a much more contaminating behaviour than the first group, we would obtain the following set of model equations:

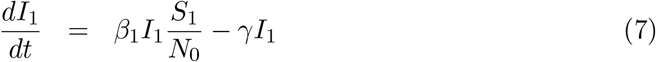

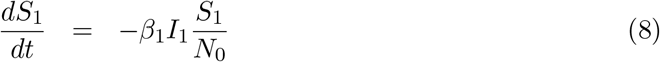

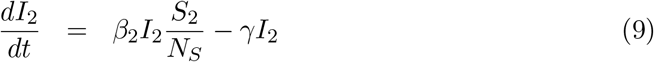

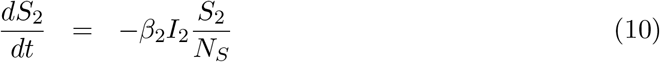

The recovery time for both contaminating groups, which is described by *γ*, is of course the same, but the first group has a coefficient of daily potential contaminations equal to *β*_1_ and the second group has a much higher *β*_2_ number of potential daily contaminations.

It isn’t very interesting to study this model, because they are simply two independent SIR models. It becomes more interesting if we introduce the fact that our super spreaders do contaminate also a lot of people from the first group:

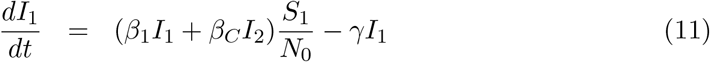

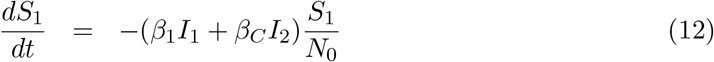

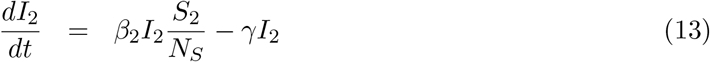

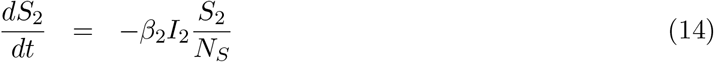

The difference with the previous set of equations resides in the extra term *β_C_I*_2_, added to *β*_1_*I*_1_ in the first two equations. It means that each super spreader (in the box *I*_2_) will on average contaminate, on top of his colleagues from group 2, potentially also *β_C_* people from group 1 per day. So people from group 1 can be contaminated by other people from group 1, or from super spreaders from group 2.

### 2.3 Example runs

#### 2.3.1 Base model

We propose the following values of the parameters for our first model, Model 1, which will serve as baseline:

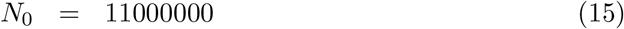

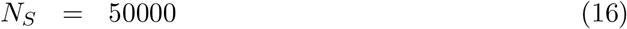

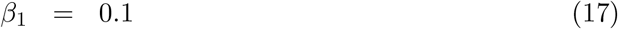

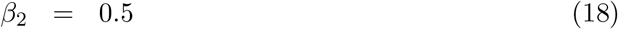

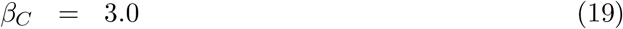

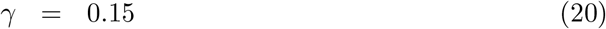

The size of the population is like the population of Belgium, with a super spreader cohort of about 50 000 people. In the general population, 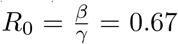 which means that the virus doesn’t propagate epidemically in the general population. However, in the spreader cohort, *R*_0_ = 3.33. And a spreader contaminates 3 people per day in the general population. The average time that one is contagious equals 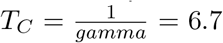 days. We start out with 5 people contaminated in the general population, but 50 people in the spreader cohort on day 0.

**Figure 4:**
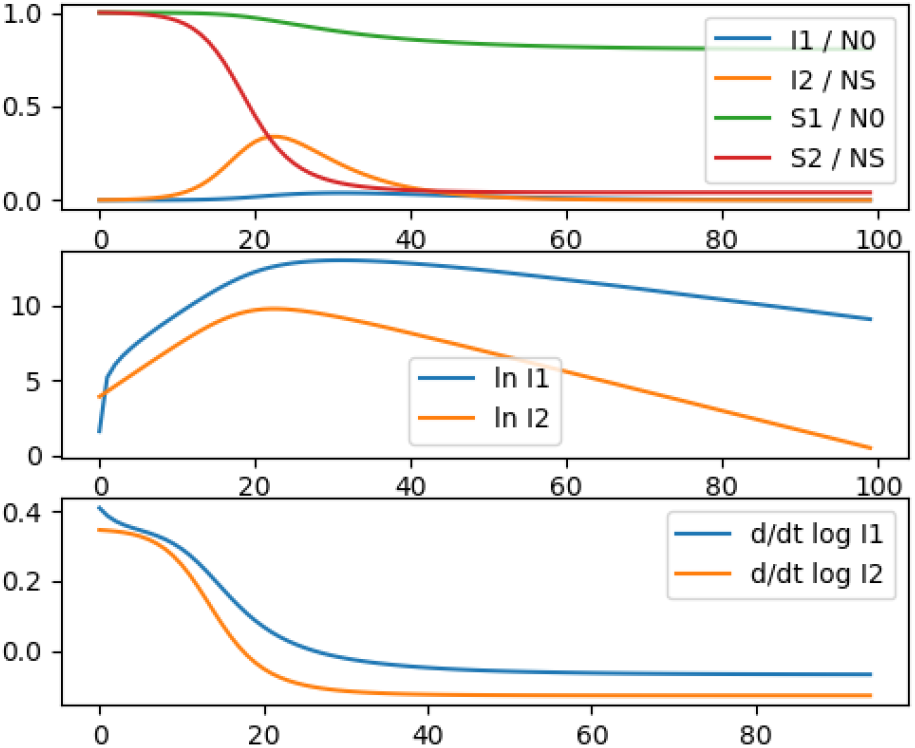
Coupled model calculation: Model 1

The spreader cohort simply follows the laws of a simple SIR model, and reaches rather quickly herd immunity and beyond (more than 95% of the spreader cohort ends up infected). The spreader cohort serves also as a driver for the dynamics in the general population, and the general population logarithmic derivative follows the spreader cohort logarithmic derivative. The slowdown of the epidemic is slower in the general population than it was in the super spreader cohort because *β*_2_*S*_2_/*N_S_* ends up being smaller than *β*_1_*S*_1_/*N*_0_. As such, it seems that the epidemic is still “keeping on” for a long time in the population. At the same time, the speed of infection slows down significantly after 2 weeks while only 5% of the population is infected, which seems low as compared to the estimated necessary herd immunity if one interprets this dynamics in the frame of a simple SIR model. In the end, about 20% of the population will end up having been infected.

#### 2.3.2 Model 2: influence of *β*_1_

If we increase now *β*_1_ to 0.14, keeping all the rest equal, so we bring the general population “closer to criticality”, and we call this Model 2, we obtain:

**Figure 5:**
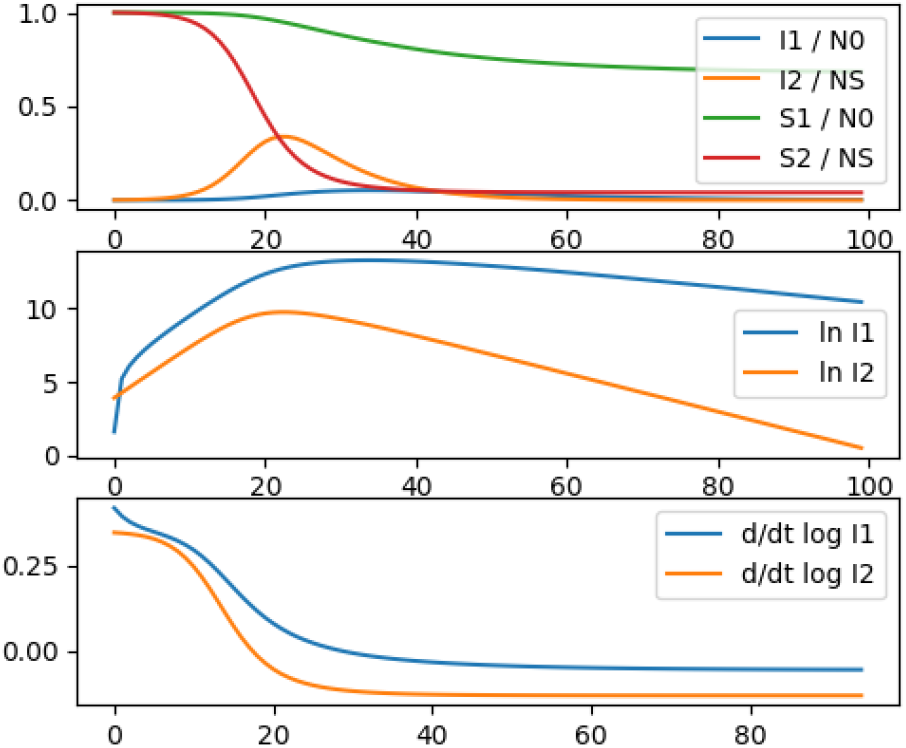
Coupled model calculation: Model 2

The dynamics is very similar to the base model, except that we now have about 32 % of the general population that will get infected. We see that bringing the population close to criticality (*R*_0_ close to 1) can significantly increase the amount of people that get infected. However, lowering *β*_1_ instead of increasing it, down to 0.03 from the 0.1 of the base model, will reduce the amount of infected people only moderately (we will obtain about 10 % of the people infected). In order to reduce *β*_1_ from 1 down to 0.03 takes a significant effort for a non-proportional gain in infected people.

#### 2.3.3 Model 3: influence of *β*_C_ and *β*_2_

Reducing the coupling *β_C_* from 3.0 down to 1.0 will have a larger effect on the people involved: only 7.7 % gets infected.

Trying to reduce the self-infection rate in the super spreaders (bringing *β*_2_ down from 0.5 to 0.3) will not reduce the end result much in the general population, but will spread out the epidemic quite longer in time. We call this last modification: Model 3.

**Figure 6:**
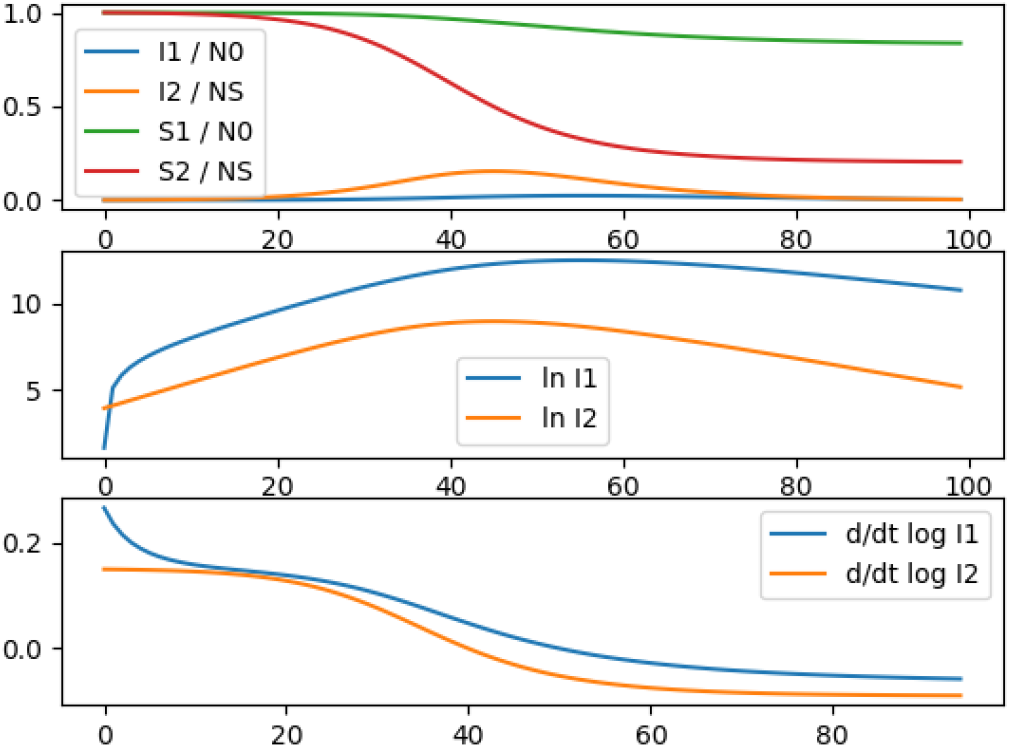
Coupled model calculation: Model 3

As we can see, the general population will end up still with 16 % infected cases, but everything is much more spread out in time.

#### 2.3.4 Model 4: supercritical general population

Let us look at a case where the illness is epidemic in the general population, be it at a very low level. That is to say, let us consider an *R*_0_ slightly above 1. We put *β*_1_ at 0.2 (so slightly larger than *γ* which is 0.15), which gives us an *R*_0_ value of 1.33. Herd immunity corresponds to 25% of the population. We also diminish the coupling of our super spreaders to the general population to a much lower value by putting *β_C_* to 0.5. We call this Model 4.

**Figure 7:**
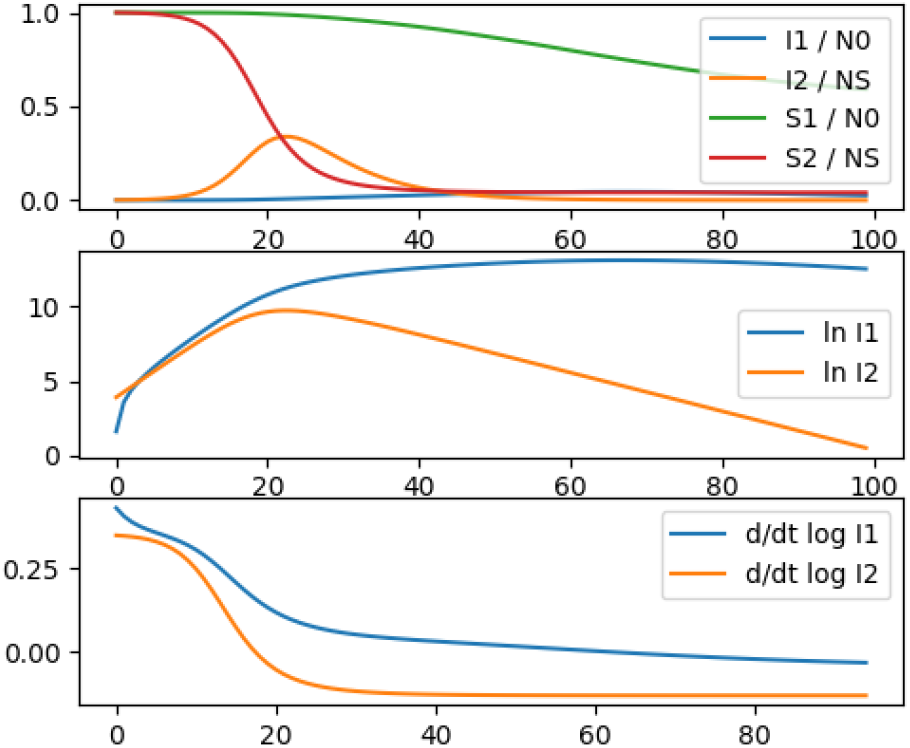
Coupled model calculation: Model 4

As we can see, the behaviour is still very similar to the behaviour of the base model. However, this time, the logarithmic derivative remains a very long time slightly above 0. It is remarkable that even with a very slight coupling between the super spreaders and the general population, the initial logarithmic derivative of the general population is still dominated by the influence of the super spreaders.

## 3 Discussion

With a simple coupled model of a small super spreader population within a general population, we can obtain the qualitative characteristics of the a priori puzzling evolution of the logarithmic derivative of proxies of the covid-19 epidemic evolution as found in several countries. Indeed, the dynamics of this epidemic seems paradoxical at first sight, when interpreted in the frame of a simple SIR model: the very high apparent *R*_0_ derived from the extremely fast doubling times of the epidemic would hint at a huge peak, and a necessary herd immunity which would be above 60% of the population. Given the CFR of the order of 1%, that has lead to predictions of large numbers of death. But the epidemic seems to have a quickly diminishing logarithmic derivative of most of its proxies, be it hospital admissions or deceased patients, while at the same time absolutely not reaching anything near herd immunity. Of course, one may think that this is due to the policies put into place. But the differences in policies put in place, and the universality of this observed dynamics may raise questions as to the influence of these policies versus the dynamics of this epidemic.

The coupling between a super spreader group that does reach herd immunity and the larger population can qualitatively explain such a phenomenon. If the covid-19 dynamics has such a kind of phenomenon as a basis, then the dynamics seems to be quite independent of any non-pharmaceutical measures taken.

The model calculations show that even if the policies do have influences on the *β* parameters, these influences don’t modify strongly the overall behaviour. If the overall population had already an *R*_0_ significantly smaller than 1, then all policies decreasing this value even further don’t have much effect on the outcome: it is only if the initial *R*_0_ is very close to 1 that they might have a effect. If measures diminish the coupling *β_C_* between the overall population and the super spreader cohort, then the effect will be significant, though. Finally if the measures diminish *β*_2_ in the spreader cohort, this will only spread out the whole dynamics in time, but it will not affect much the end result.

This may explain why there seems to be a universal behaviour to the epidemic, quite insensitive to specific policies that try to diminish the *β* factors without having a very strong effect on *β_C_*, the only parameter that seems to have a proportional influence on the outcome.

The coupled dynamics of a super spreader cohort within a general population that isn’t critical is quite robust against non-pharmaceutical measures. But even in a general population that is slightly critical, a small group of super spreaders will dominate the behaviour in the beginning of the epidemic. The only difference is that the value of the logarithmic derivative is slightly positive after a while, instead of taking on a negative value. It might be that Brazil is in this case.

## 4 Conclusion

The model of a population in which the disease cannot propagate epidemically or would give rise to a very slightly propagating epidemic with an *R*_0_ value near one, in which a strongly coupled, but small, super spreader cohort is present in which the disease is strongly epidemic (large *R*_0_ value in that group) will have a robust dynamics that resembles the one observed of the covid-19 pandemic, displaying a very high initial logarithmic derivatives, decreasing steadily towards a small value. This dynamics is quite robust against non-pharmaceutical measures that modify the individual *β* values of this model.

## Data Availability

All data is public and all links are in the paper as references.

